# Incidence of and Patient-Reported Reasons for Transfer-Out Among People Receiving Long-Acting Antiretroviral Therapy in Ndola District, Zambia: A Multisite Cross-Sectional Study of Formal and Silent Transfers

**DOI:** 10.64898/2026.11.18.26368457

**Authors:** MacMillan Mwale, Mwiya Mombotwa, Siame Lukundo

## Abstract

**Background:** Transfer-out is a common outcome in HIV treatment programmes, and transfers that are not documented (“silent transfers”) are often misclassified as loss to follow up. Understanding why recipients of care move between facilities, and which reasons lead to undocumented moves, can guide retention strategies. We estimated the incidence of transfer-out and described patient-reported reasons for formal and silent transfer among people receiving antiretroviral therapy (ART) in Ndola District, Zambia.

**Methods:** We conducted a cross-sectional study in 11 ART facilities between April and October 2026. All recipients of care who transferred during the study period and consented were interviewed using a World Health Organization (WHO)-adapted questionnaire: formal transfers at exit and silent transfers by telephone once re-engagement elsewhere was established. Reasons were coded into structural, psychosocial and clinic-based domains. We report proportions with Wilson 95% confidence intervals (CIs) and used Firth penalized logistic regression to identify factors associated with silent rather than formal transfer.

**Results:** Of 9,866 recipients of care on ART, 120 transferred out (cumulative incidence 1.22%; 95% CI 1.02–1.45%). The median age was 41 years (IQR 32–48.5) and 61.3% were female; 12 (10.0%) were silent transfers. Structural reasons accounted for 90.8% (95% CI 84.3–94.8) of transfers: work, education or business (55.0%), relocation (32.5%) and distance or transport (3.3%). Psychosocial reasons (marriage or family obligations) accounted for 9.2% (95% CI 5.2–15.7), and no participant reported a clinic-based reason. Silent transfer was more frequent among those reporting psychosocial than structural reasons (45.5% vs 6.4%; adjusted odds ratio [aOR] 11.36 (2.75–49.56)). Silent transfers were younger than formal transfers (median 33 vs 41 years; P = 0.031), while sex was not associated with transfer type.

**Conclusions:** Transfer-out in Ndola District was uncommon and driven almost entirely by mobility linked to work and relocation rather than dissatisfaction with care. Transfers prompted by marriage or family obligations were much more likely to be undocumented. Asking recipients of care about upcoming life changes, offering facilitated transfers, and using a shared electronic health record could keep mobile recipients of care visible in the HIV programme.

## Introduction

Retention in care is essential for people living with HIV to achieve and sustain viral suppression, and differentiated service delivery models aim to make lifelong ART more convenient [1]. However, attrition remains substantial in sub-Saharan Africa and increases with time on treatment [2]. Some patients classified as lost to follow-up (LTFU) have instead transferred to other facilities without notifying their original clinics. An individual-patient-data meta-analysis across eight African countries found that 14.8% of patients classified as LTFU had transferred elsewhere [3], while South African national laboratory data showed that accounting for inter-facility movement substantially improves retention estimates [4].

Transfers may be formal, with the receiving facility documented, or undocumented (silent transfers). Silent transfers inflate attrition estimates, misdirect tracing resources and distort programme indicators through duplicate registration [5,9,12]. They may also interrupt treatment when patients encounter administrative barriers at receiving facilities [13,14]. This has clinical implications: studies in South Africa and Uganda have associated inter-facility transfers with viraemia and viral rebound [10,15,16]. In Zambia, 26% of patients classified as LTFU who were traced alive had moved to another clinic without documentation; only a quarter of verified transfers involved official paperwork, and the median gap between visits at the two facilities was 188 days [7].

Understanding the reasons for clinic transfers is important for improving retention strategies. In Zambia, Sikazwe et al. classified reasons for clinic switching into structural (access-related), psychosocial (personal) and clinic-based (service delivery) barriers [6]. Structural barriers predominated among silent transfers, particularly relocation (35.5%), work obligations (29.7%) and easier or cheaper transport (27.2%). Barrier type was also associated with whether patients re-engaged elsewhere or disengaged from care [6]. South African studies similarly identified mobility as the main reason for clinic transfers, with younger age also associated with transfer [9,10].

However, existing studies have largely examined patients already classified as LTFU, leaving limited evidence on transfer-out incidence and determinants of silent transfers among recipients of care in routine Zambian settings. We therefore estimated transfer-out incidence, described patient-reported reasons and examined factors associated with silent versus formal transfer among recipients of care attending 11 ART facilities in Ndola District, an urban setting characterised by labour and residential mobility.

## Methods

### Study design and setting

We conducted a multisite cross-sectional study in 11 public ART facilities in Ndola District, Copperbelt Province, Zambia, between April and October 2026. Facilities were purposively selected on the basis of having a data associate able to complete the data-capture tool and a peer navigator able to conduct participant interviews. HIV care at these facilities follows the Zambia Consolidated Guidelines for Treatment and Prevention of HIV Infection [11].

### Participants and definitions

All recipients of care receiving ART at a participating facility who transferred out during the study period and consented were eligible; no sampling within facilities was performed. We defined a **formal transfer** as a transfer requested by the recipient of care and documented by the sending facility, with the receiving facility known, and a **silent transfer** as a recipient of care who stopped attending the sending facility without a documented transfer and was subsequently established to be receiving ART at another facility [6,7]. Silent transfers were identified via missed-appointment tracing, phone calls or confirmation from the receiving facility. The denominator for incidence was the number of recipients of care currently on treatment (TX_CURR) at the 11 facilities [state reporting date/quarter].

### Procedures and measurements

Peer navigators administered a structured questionnaire adapted from WHO patient-monitoring tools. Recipients of care undergoing formal transfer were interviewed at exit; silent transfers were interviewed by telephone once their transfer status had been established. The questionnaire recorded age, sex, type of transfer, the main reason for transfer and the destination facility. Interviews were conducted in English or a local language as preferred by the participant.

### Coding of reasons

We coded each reported reason into one of three domains: **structural barriers**, covering changes in circumstance or access, including relocation, work, education or business obligations, and distance or transport to the clinic; **psychosocial barriers**, covering personal and social circumstances, including marriage and other family obligations, stigma or fear of disclosure; and **clinic-based barriers**, covering service-delivery factors such as waiting time, staff attitudes and perceived quality of care [6]. Free-text responses were recorded into these categories by coder(s)with disagreements resolved by discussion.

### Statistical analysis

Data were entered into Microsoft Excel, cleaned and checked for consistency. Non-standard entries were harmonised (for example, spelling variants of “informed”; records labelled “patient request” or “relocated” in the transfer-type field were classified as formal transfers). We calculated the cumulative incidence of transfer-out as the number of recipients of care who transferred out divided by TX_CURR, with a Wilson 95% CI. We summarised participant characteristics as frequencies and percentages or medians with interquartile ranges (IQR), overall and by transfer type, and compared formal and silent transfers using Fisher’s exact test for sex, a Monte Carlo permutation test (20,000 permutations) for age group and reason category, and the Mann–Whitney U test for age.

We report the proportion of participants citing each reason and domain with Wilson 95% CIs, overall and by transfer type. To examine factors associated with silent rather than formal transfer, we fitted logistic regression models with sex, age (per 10 years) and reason domain (psychosocial vs structural). Because silent transfers were few (11 events among 119 participants with complete data), we used Firth penalized likelihood to reduce small-sample bias and report odds ratios (ORs) with profile penalized-likelihood 95% CIs. The regression was considered exploratory. Participants with missing age or sex (n = 1) were excluded from the age, sex and regression analyses. Analyses were performed in Python 3.13 (statsmodels and SciPy).

### Ethical considerations

The study was approved by Tropical Diseases Research Center Ethics committee and permission was obtained from Ndola District Health Office / Provincial Health Office. Written informed consent was obtained from adult participants before interview; for participants younger than 18 years, the guardians gave consent and participant assented. Verbal consent was obtained and documented for telephone interviews. No personal identifiers were entered into the analysis dataset, and participation did not affect the care received.

## Results

### Participants and incidence of transfer-out

Across the 11 facilities, 9,866 recipients of care were on ART, 140 transfers occurred and 120 were interviewed, report incidence as 140/9,866 = 1.42% (95% CI 1.20–1.67%) and a response rate of 85.7%. Of the 120 participants, 108 (90.0%; 95% CI 83.3–94.2%) were formal transfers and 12 (10.0%; 95% CI 5.8–16.7%) were silent transfers. (Figure 1).

**Figure 1.**
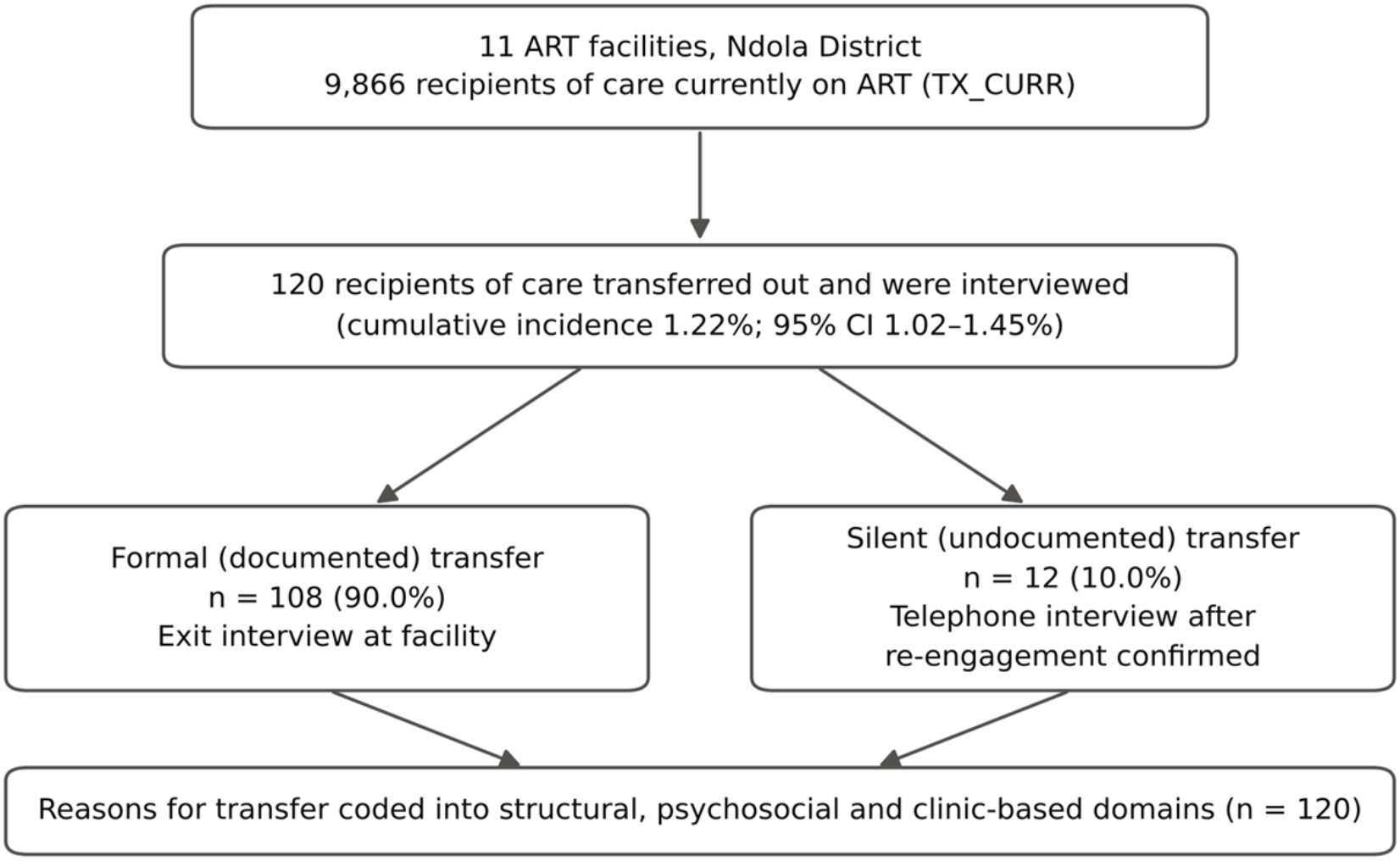
Flow of recipients of care through the study. CI, confidence interval; TX_CURR, number of recipients of care currently on treatment.

The median age was 41 years (IQR 32–48.5; range 11–61) and 73 of 119 participants with recorded sex (61.3%) were female (Table 1). Silent transfers were younger than formal transfers (median 33 vs 41 years; P = 0.031): 6 of 11 silent transfers with recorded age (54.5%) were aged 25–34 years compared with 20.4% of formal transfers, and only one (9.1%) was aged 45 years or older compared with 38.9% of formal transfers. The proportion female was similar in both groups (63.6% vs 61.1%; P = 1.000).

**Table 1.** Characteristics of recipients of care who transferred out, by transfer type, Ndola District, Zambia, 2026.

| **Characteristic** | **All (N = 120)** | **Formal transfer (n = 108)** | **Silent transfer (n = 12)** | **P value** |
|---|---|---|---|---|
| **Sex** |  |  |  |  |
| Female | 73 (61.3) | 66 (61.1) | 7 (63.6) | 1.000<sup>a</sup> |
| Male | 46 (38.7) | 42 (38.9) | 4 (36.4) |  |
| **Age, years** |  |  |  |  |
| Median (IQR) | 41 (32–48.5) | 41 (33–49) | 33 (28.5–40) | 0.031<sup>b</sup> |
| Range | 11–61 |  |  |  |
| <25 | 11 (9.2) | 10 (9.3) | 1 (9.1) | 0.048<sup>c</sup> |
| 25–34 | 28 (23.5) | 22 (20.4) | 6 (54.5) |  |
| 35–44 | 37 (31.1) | 34 (31.5) | 3 (27.3) |  |
| ≥45 | 43 (36.1) | 42 (38.9) | 1 (9.1) |  |

Data are n (%) unless stated otherwise. Percentages exclude missing values. IQR, interquartile range.
<sup>a</sup> Fisher's exact test. <sup>b</sup> Mann–Whitney U test. <sup>c</sup> Monte Carlo permutation test of the chi-square statistic (20,000 permutations).

### Reasons for transfer-out

Structural barriers accounted for 109 transfers (90.8% (95% CI 84.3–94.8)) (Table 2, Figure 2). Employment and work obligations were the single most common reason (53.3% (95% CI 44.4–62.0)); combined with education (n = 1) and business activities (n = 1), work-, study- or business-related mobility accounted for 66 transfers (55.0%; 95% CI 46.1–63.6%). Relocation to a new area was cited by 39 participants (32.5%; 95% CI 24.8–41.3%) and distance or transport to the clinic by 4 (3.3%; 95% CI 1.3–8.3%).

**Table 2.** Patient-reported reasons for transfer-out, by domain and transfer type (N = 120)

| **Domain and reason** | **n** | **% of all (95% CI)** | **Formal, n (%) (n = 108)** | **Silent, n (%) (n = 12)** |
|---|---|---|---|---|
| **Structural (access) barriers** | **109** | **90.8 (84.3–94.8)** | **102 (94.4)** | **7 (58.3)** |
| Employment / work obligations | 64 | 53.3 (44.4–62.0) | 59 (54.6) | 5 (41.7) |
| Relocated to a new area | 39 | 32.5 (24.8–41.3) | 38 (35.2) | 1 (8.3) |
| Distance / transport to clinic | 4 | 3.3 (1.3–8.3) | 3 (2.8) | 1 (8.3) |
| Education (school/college) | 1 | 0.8 (0.1–4.6) | 1 (0.9) | 0 (0.0) |
| Business activities | 1 | 0.8 (0.1–4.6) | 1 (0.9) | 0 (0.0) |
| **Psychosocial barriers** | **11** | **9.2 (5.2–15.7)** | **6 (5.6)** | **5 (41.7)** |
| Marriage / family obligations | 11 | 9.2 (5.2–15.7) | 6 (5.6) | 5 (41.7) |
| **Clinic-based barriers** | **0** | **0.0 (0.0–3.1)** | **0 (0.0)** | **0 (0.0)** |
| Waiting time, staff attitude, quality of care | 0 | 0.0 (0.0–3.1) | 0 (0.0) | 0 (0.0) |

Domains follow the framework of Sikazwe et al. [6]. CIs are Wilson score intervals. Percentages in the formal and silent columns use column totals as denominators.

**Figure 2.**
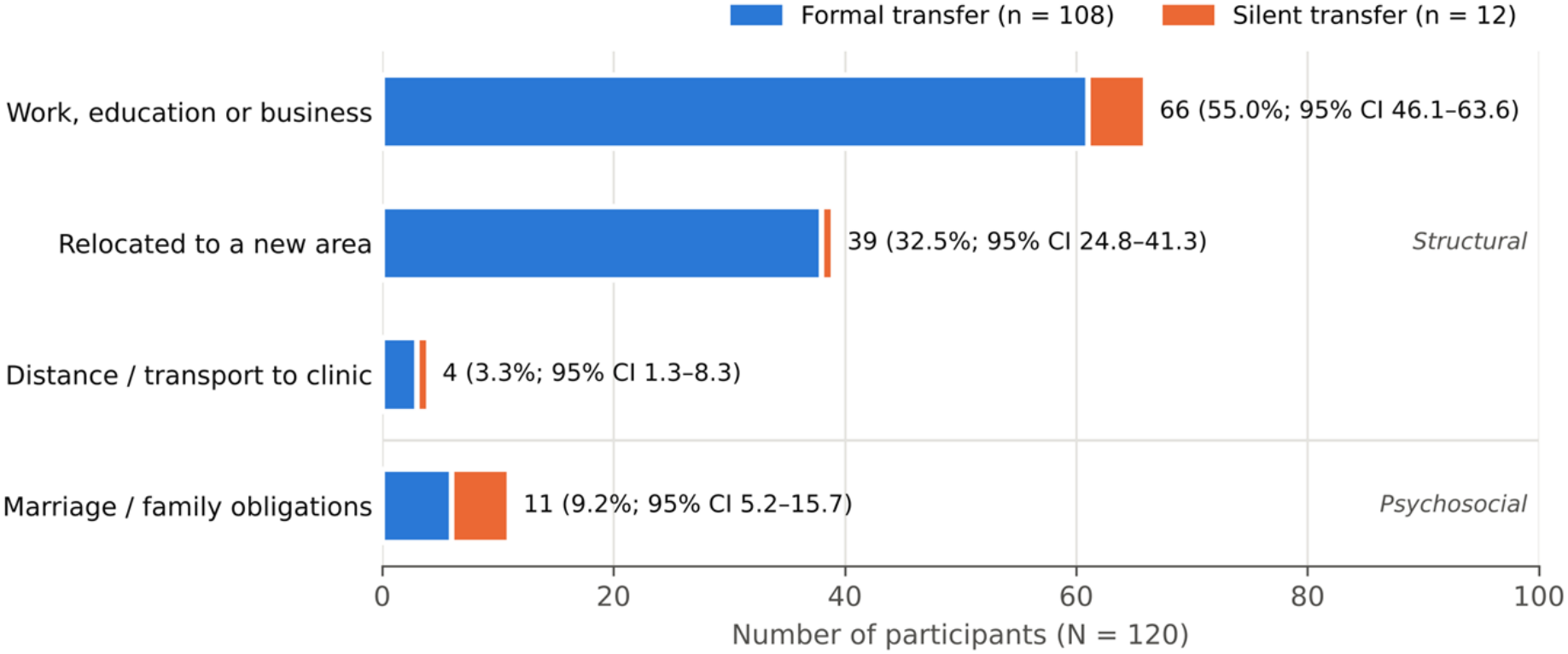
Main reason for transfer-out among recipients of care, split by formal and silent transfer. Labels show the total number, the percentage of all participants and the Wilson 95% confidence interval. No participant reported a clinic-based reason.

Psychosocial barriers, all in the form of marriage or other family obligations, accounted for 11 transfers (9.2% (95% CI 5.2–15.7)). No participant reported a clinic-based reason such as long waiting times, staff attitudes or quality of care (0%; 95% CI 0.0–3.1%). Reason domain did not differ by sex: psychosocial reasons were reported by 6 of 46 men (13.0%) and 5 of 73 women (6.8%; P = 0.333). Each participant reported a single main reason, so overlap between domains could not be assessed.

### Factors associated with silent transfer

The share of transfers that were silent differed by reason (P = 0.003, permutation test). Five of the 11 transfers driven by marriage or family obligations (45.5%) were silent, compared with 5 of 66 driven by work, education or business (7.6%), 1 of 39 driven by relocation (2.6%) and 1 of 4 driven by distance (25.0%). In Firth logistic regression, a psychosocial reason was strongly associated with silent rather than formal transfer (crude OR 13.34 (3.32–55.47); adjusted OR 11.36 (2.75–49.56); Table 3). Older age showed a trend toward lower odds of silent transfer (aOR per 10 years 0.63 (0.33–1.17)), and sex was not associated with transfer type (aOR 0.91 (0.20–3.69)). Including the participant with missing age and sex in a model with reason domain alone gave a similar estimate (OR 11.56 (2.94–46.35)).

**Table 3.**
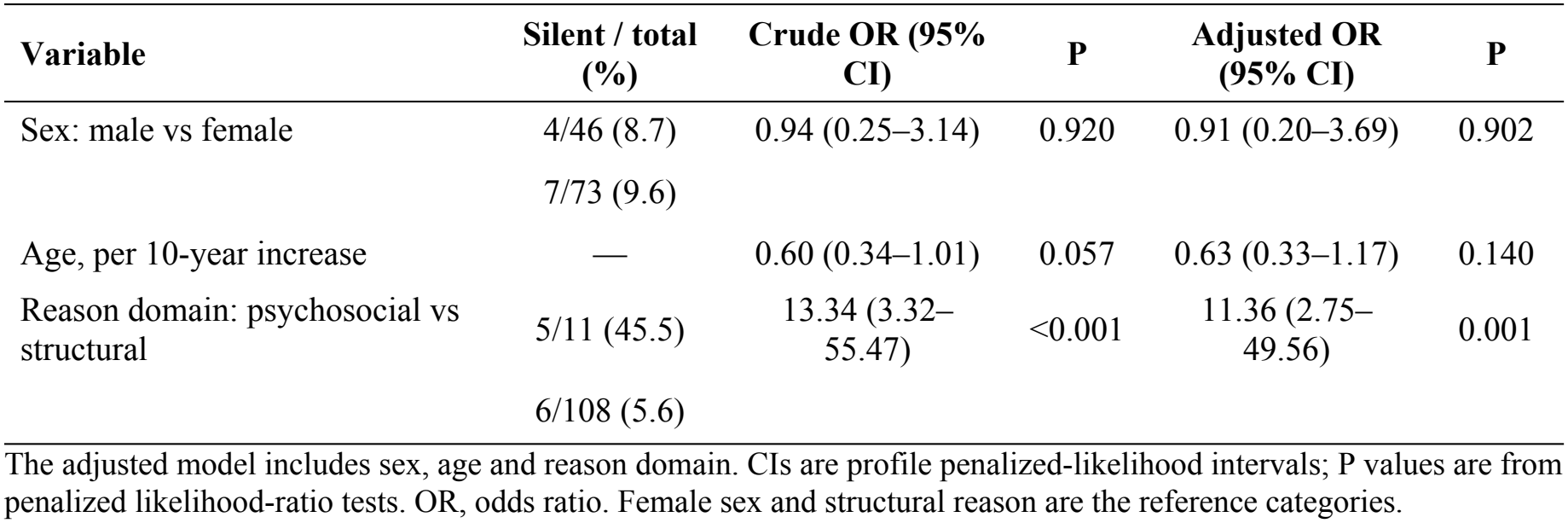
Factors associated with silent rather than formal transfer: Firth penalized logistic regression (n = 119; 11 silent transfers)

| **Variable** | **Silent / total (%)** | **Crude OR (95% CI)** | **P** | **Adjusted OR (95% CI)** | **P** |
|---|---|---|---|---|---|
| Sex: male vs female | 4/46 (8.7) | 0.94 (0.25–3.14) | 0.920 | 0.91 (0.20–3.69) | 0.902 |
|  | 7/73 (9.6) |  |  |  |  |
| Age, per 10-year increase | — | 0.60 (0.34–1.01) | 0.057 | 0.63 (0.33–1.17) | 0.140 |
| Reason domain: psychosocial vs structural | 5/11 (45.5) | 13.34 (3.32–55.47) | <0.001 | 11.36 (2.75–49.56) | 0.001 |
|  | 6/108 (5.6) |  |  |  |  |

The adjusted model includes sex, age and reason domain. CIs are profile penalized-likelihood intervals; P values are from penalized likelihood-ratio tests. OR, odds ratio. Female sex and structural reason are the reference categories.

## Discussion

In this multisite study in Ndola District, about 1 in 80 recipients of care on ART transferred out during the study period, and nine in ten of those transfers were driven by structural reasons linked to mobility: work, education or business, and relocation. No participant cited dissatisfaction with the clinic. One in ten transfers was silent, and silent transfers were concentrated among recipients of care who moved because of marriage or family obligations and among younger adults. These findings suggest that transfer-out in this urban setting largely reflects the normal movement of working-age people, and that the risk of falling out of view of the programme depends on why people move.

The predominance of structural reasons closely mirrors the Zambian estimates of Sikazwe et al., in which relocation, work obligations and transport were the leading reasons for silent transfer [6], and South African cohorts in which mobility explained about three-quarters of clinic transfers [9]. Our finding that work, education and business were the single largest category (55.0%) is consistent with Ndola’s role as an industrial and commercial centre, where job changes, postings and seasonal work are common. The absence of clinic-based reasons differs from Sikazwe et al., where 13.3% of silent transfers cited shorter clinic visits elsewhere [6]. This may reflect genuinely acceptable services in the participating facilities, including differentiated service delivery and multi-month dispensing [1], but may also reflect social desirability bias, since most interviews were conducted at exit by peer navigators attached to the sending facility.

The proportion of silent transfers (10.0%) was much lower than in the tracing studies of Sikombe et al., in which only about a quarter of verified transfers carried official paperwork [7]. The two estimates measure different things: Sikombe et al. traced patients already classified as lost, whereas we enumerated transfers prospectively at the sending facility, where formal transfers are fully visible and silent transfers can only be counted once they are found. Our figure is therefore best interpreted as a minimum, and the true share of undocumented moves in Ndola is probably higher.

The strongest finding was the association between psychosocial reasons and silent transfer: nearly half of transfers prompted by marriage or family obligations were undocumented, compared with about 6% of those prompted by structural reasons. Sikazwe et al. likewise found that the type of barrier, rather than demographic characteristics, was what distinguished patients who moved to a new clinic from those who left care, and that psychosocial barriers marked those at greatest risk of falling out of care [6]. Marriage-related moves can be sudden, may involve joining a new household in which HIV status has not been disclosed, and may lead recipients of care to avoid asking for a transfer letter. Although our numbers were small and the CI is wide, this suggests a group in whom transfer counselling and attention to disclosure could prevent unrecorded moves. Qualitative work among postpartum women in Cape Town identified similar drivers: conflict with partners or family, fear of disclosure and community stigma prompted moves, and silent transfers were often linked to poor relationships with health workers [13].

Silent transfers were also younger than formal transfers. Younger age has been associated with transfer-out in South African cohorts [9,10] and with disengagement more broadly [2], and younger adults are the most mobile for work, study and marriage. In a population-based Ugandan cohort, facility switching was likewise most frequent among people younger than 30 years [16], and age under 30 was independently associated with transfer in the Western Cape [15]. The comparable sex distribution in formal and silent transfers contrasts with the lower re-engagement observed among men in Zambia [6], but our study included only recipients of care who had re-engaged and were reachable, so men who left care entirely would not have been captured.

The cumulative incidence of 1.22% is lower than the 4.7 per 100 person-years reported in Cape Town [10] and the 19% of patients who transferred out over about two and a half years in Mzuzu, Malawi [5]. Because our estimate covers only a few months and uses a cross-sectional TX_CURR denominator rather than person-time, these figures are not directly comparable, and an annualized figure would be roughly twice as high. Even so, transfer-out appears to be a small but persistent flow out of each facility, and its accurate recording matters for retention indicators [4]. A population-based cohort in Rakai, Uganda, reported facility switching at 4.8 per 100 person-years over a longer follow-up period [16].

### Implications

Three practical implications follow. First, because most transfers are predictable consequences of job changes and moves, facilities can routinely ask recipients of care at each visit about upcoming relocations and offer a facilitated transfer, with a referral letter, a confirmed receiving facility and enough medication to cover the gap. Second, people transferring because of marriage or family obligations warrant particular attention, including counselling on disclosure and on how to continue care discreetly at the new location. Third, a shared electronic health record with a unique ART identifier would allow the receiving facility to see prior treatment history and the sending facility to reclassify silent transfers; in Zambia, keeping the same ART number at the new clinic was associated with faster resumption of treatment [7]. Clinical follow-up after transfer also deserves attention: transfer has been associated with viraemia in South Africa [15] and with viral rebound in Uganda [16]. A review of South African guidelines found that the clinical management of people transferring between primary care facilities was not specified [12]; Zambian transfer procedures could explicitly include an adherence review and an early viral load at the receiving facility.

### Strengths and limitations

Strengths of this study include its multisite design, the prospective inclusion of transfers at the point they occurred, the inclusion of silent transfers through telephone follow-up, and the use of a published framework that allows direct comparison with national Zambian data [6]. The study also has limitations. Facilities were purposively selected and participation depended on consent and reachability, so results may not generalise to the whole district. Silent transfers who could not be found were not included, and their number is therefore underestimated. Only a single main reason was recorded, whereas most patients in Sikazwe et al. reported several [6]; this may have hidden secondary psychosocial or clinic-based reasons. We had no comparison group of recipients of care who did not transfer, so we could describe reasons but not identify risk factors for transfer-out itself. Facility identifiers were not recorded, so we could not examine variation between facilities, which Sikazwe et al. found to be substantial [6]. Clinical variables such as time on ART, viral load and differentiated service delivery model were not collected. Finally, the small number of silent transfers produced wide CIs, and the regression results should be regarded as exploratory.

### Conclusions

Transfer-out among recipients of care on ART in Ndola District was uncommon and almost entirely driven by structural mobility linked to work and relocation, not by dissatisfaction with services. Transfers driven by marriage or family obligations, and those among younger adults, were more likely to be undocumented. Asking recipients of care about upcoming moves, offering facilitated transfers with tailored counselling, and linking facilities through a shared electronic record could help mobile recipients of care stay visible in, and connected to, HIV care.

## Data Availability

Data available on request.

## Acknowledgements

We would like to acknowledge facility staff, data associates, peer navigators and participants.

## Declarations

### Data availability

The de-identified dataset can be obtained.

### Author contributions

Dr. MacMillan Mwale-Principal Investigator, Dr. Siame Lukundo-Data Analysis and report editing and Mrs. Mwiya Mombotwa-Study logistics and Data collection.

